# Transcriptome-wide Association Study of Circulating IgE Levels Identifies Novel Targets for Asthma and Allergic Diseases

**DOI:** 10.1101/2020.08.17.20176479

**Authors:** Recto Kathryn, Huan Tianxiao, Lee Dong Heon, Lee Gha Young, Gereige Jessica, Yao Chen, Hwang Shih-Jen, Joehanes Roby, Rachel S Kelly, Lasky-Su Jessica, O’Connor George, Levy Daniel

## Abstract

Measurement of circulating immunoglobulin E (IgE) concentration is helpful for diagnosing and treating asthma and allergic diseases. Identifying gene expression signatures associated with IgE might elucidate novel pathways for IgE regulation. To this end, we performed a discovery transcriptome-wide association study (TWAS) to identify differentially expressed genes associated with circulating IgE levels in whole-blood derived RNA from 5,345 participants in the Framingham Heart Study (FHS) across 17,873 mRNA gene-level transcripts. We identified 216 significant transcripts at a false discovery rate (FDR)< 0.05. We conducted replication using the meta-analysis of two independent external studies: the Childhood Asthma Management Program (n = 610) and the Genetic Epidemiology of Asthma in Costa Rica Study (n = 326); we then reversed the discovery and replication cohorts, which revealed 59 significant genes that bidirectionally replicated. Gene ontology analysis revealed that many of these genes were implicated in immune function pathways, including defense response, inflammatory response, and cytokine production. Mendelian randomization (MR) analysis revealed four genes (*CLC, CCDC21, S100A13*, and *GCNT1*) as putatively causal (p< 0.05) regulators of IgE levels. *GCNT1* (beta = 1.5, p = 0.01)—which is a top result in the MR analysis of expression in relation to asthma and allergic diseases—plays a role in regulating T helper type 1 (Th1) cell homing, lymphocyte trafficking, and B cell differentiation. Our findings build upon prior knowledge of IgE regulation and provide a deeper understanding of underlying molecular mechanisms. The IgE-associated genes that we identified—particularly those implicated in MR analysis—can be explored as promising therapeutic targets for asthma and IgE-related diseases.

## Introduction

Immunoglobulin E (IgE) is an antibody produced by B cells located in lymph nodes in response to antigenic stimuli and its production requires T helper type 2 (Th2) cells^1^. Once released into the circulation, IgE contributes to immunity to respiratory viruses and parasites and protects against venom toxin exposure^2,3^. IgE also plays a role in disease processes related to allergic asthma, allergic rhinitis, atopic dermatitis, and food allergies^4^. According to recent estimates from the World Health Organization, asthma affected 300 million people worldwide in 2012 and this number is projected to increase to 400 million by 2025^4^. Given the widespread burden of IgE-mediated allergic diseases, investigating the maladaptive role of IgE in immune responses may highlight promising therapies for asthma and related conditions.

Genome-wide association studies (GWAS) have identified single nucleotide polymorphisms (SNPs) at the *STAT6, FCER1A, IL13, IL4/RAD50*, and the major histocompatibility complex (MHC) loci that are associated with circulating IgE concentrations^5–8^. Investigating the transcriptomic signature of IgE concentration may shed light on molecular regulatory mechanisms^9–11^. Virkud et al. examined gene expression networks in whole-blood in two differing asthma populations and identified 31 transcripts associated with serum total IgE that replicated across study cohorts^12^. To date, however, there have been no published large-scale transcriptome-wide association studies (TWAS) of circulating IgE concentration. While most of the current literature has focused on certain aspects of IgE-related gene regulatory networks, our study was designed to provide a more comprehensive framework for understanding the molecular regulation of IgE by integrating TWAS of IgE with GWAS of IgE and IgE-related diseases.

In this study, we hypothesized *a priori* that IgE-associated transcriptomic changes impact IgE regulation, which in turn play a role in the pathology of IgE-related diseases, such as asthma and allergic diseases. First, we performed a discovery TWAS of IgE in 5345 Framingham Heart Study (FHS) participants. To validate our results, we conducted replication based on the meta-analysis of two independent external studies: the Childhood Asthma Management Program (CAMP) and the Genetic Epidemiology of Asthma in Costa Rica Study (GACRS). We then reversed the discovery and replication sets. Second, we conducted Mendelian randomization (MR) to determine the direction of effect and infer causal relations between gene expression and circulating IgE levels. Two-sample MR analyses were then used to infer causal relations between IgE-related gene expression and IgE-related diseases, including asthma and allergy, by linking genetic variants associated with gene expression (i.e. *cis-*eQTLs) with GWAS of asthma and allergy, respectively^13^. By exploring the multidimensional interrelations of gene expression and circulating IgE levels, we provide a deeper understanding of the molecular pathways underlying IgE regulation and highlight promising therapeutic targets for IgE-related diseases.

## Methods

### Discovery in the FHS

#### Study population

A flowchart of the study design is displayed in Figure 1. The FHS is a community-based study^14^. The study sample consisted of 5345 individuals from the FHS Offspring (n = 2251) and Third Generation (n = 3094) cohorts, in whom IgE levels and gene expression were measured. The study protocol was approved by the Institutional Review Board at Boston University Medical Center (Boston, MA). All participants gave informed consent for genetic research.

**Figure 1.**
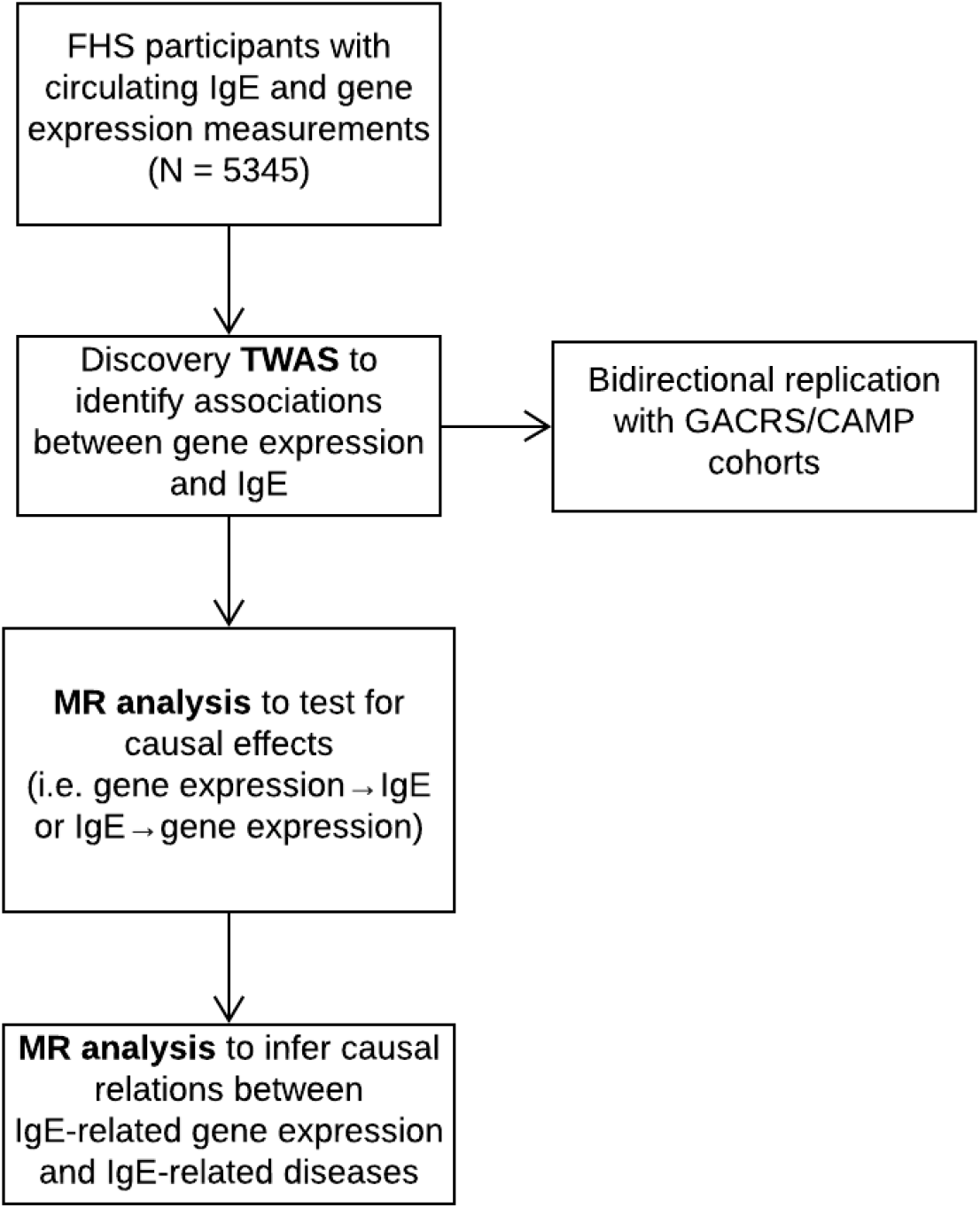
Flowchart of study design.

#### Assessment of IgE levels

,Serum total IgE concentration was measured on FHS Offspring (Exam 7: 1998–2001) and Third Generation (Exam 1: 2002–2005) cohort participants. Total IgE measurements were performed using the Phadia Immunocap 100 system, in which an anti-IgE antibody is bound to a solidphase carrier followed by fluoroenzyme-based quantitative measurement of total IgE with high precision and reproducibility^15^.

#### mRNA expression data

Gene expression was measured on FHS Offspring (Exam 8: 2005-2008) and Third Generation (Exam 2: 2008-2011) cohort participants. Whole blood samples (2.5 ml) were collected in PAXgene™ tubes (PreAnalytiX, Hombrechtikon, Switzerland). mRNA expression was profiled using the Affymetrix Human Exon 1.0 ST GeneChip (Santa Clara, CA) platform that includes 18,000 gene-level transcripts. The data normalization was described previously^16^.

#### Association of gene expression with IgE levels

A linear mixed model implemented in the *lmekin()* package in R was used to analyze associations between gene expression (RMA value) and serum total IgE concentration after adjusting for age, sex, smoking status (current, former, and never smokers), pack-years, technical covariates including batch effects^16^, predicted blood cell fraction (including white blood cells, red blood cells, platelets, lymphocytes, monocytes, and basophils), and family structure. We performed a secondary analysis further adjusting for eosinophils.

#### GWAS of IgE

There have been no large-scale GWAS of serum IgE concentration published in the past five years. Given the limited availability of up-to-date IgE GWAS, we updated a previous FHS GWAS of IgE concentration^8^ using 1000 Genomes imputation. We characterized statistical associations between genome-wide polymorphisms and variation of serum IgE concentration using a linear mixed regression model. The updated GWAS included 7252 FHS participants from three cohorts the FHS Original cohort (Exam 24; 1995-1998; n = 495), Offspring cohort (Exam 7; 1998-2001; n = 3003), and Third Generation cohort (Exam 1; 2002-2005; n = 3764). DNA samples of the FHS participants who gave consent for genomic studies were genotyped using the Affymetrix 550K array (Santa Clara, CA). We applied quality control criteria of ≥95% call rate, ≥1×10^−6^ p-value of Hardy-Weinberg equilibrium, and ≥1% minor-allele-frequency. After applying the quality-control approved genotyping, we generated imputed whole-genome polymorphism panels using the MACH platform and applied the 1000 Genomes phase 1 platform as the reference library. For the current association analysis, we tested for statistical association assuming additive influence of polymorphisms, and required an imputation quality of 20% or higher.

#### Mendelian randomization analysis

We used a two-stage least squares (2SLS) Mendelian randomization (MR) method to estimate the causal relationships between gene expression and IgE measured in 5345 FHS participants. Bi-directional MR analyses were performed to test if expression drives IgE concentration (i.e., mRNA → IgE), using the top *cis-*eQTL for each mRNA as an instrumental variable (IV)^17^, or if IgE concentration drives mRNA expression, using the genetic risk score combined by the top six loci from previous IgE GWAS results at P< 5×10^−8^ (i.e., IgE → mRNA)^5,8^. The six IgE-associated SNPs that were used in the polygenic risk score include rs2251746 (FCER1A), rs1059513 (STAT6), rs1295686 (IL13), rs2523809 (HLA-G), rs2517754 (HLA-A), and rs2858331 (HLA-DQA2)^5,8^. To determine the strength of the genetic instrument, an F-statistic in a linear regression model was derived from the proportion of variation in the exposure that was explained by the corresponding IV. *cis*-eQTLs with an F-statistic less than 10, indicating a weak instrument, were excluded. We considered an mRNA putatively causal for IgE (i.e., mRNA→IgE) when the MR test for mRNA → IgE was significant (P_mRNA→IgE_ < 0.05), and IgE → mRNA was not significant (P_IgE→mRNA_ ≥ 0.05).

Two-sample MR was used to identify putatively causal mRNAs for both asthma and allergic diseases using the MRbase package in R. Estimated associations and effect sizes between SNPs and asthma and allergic diseases were based on UK Biobank GWAS of asthma and allergic diseases (hay fever, allergic rhinitis, or eczema) phenotypes, respectively^13^. Using *cis-*eQTLs associated with gene transcripts associated with circulating IgE levels as instrumental variables, MR analyses were used to test if gene expression drives asthma/allergy (i.e., mRNA → asthma/allergy).

#### Pathway analysis

Pathway analysis using Gene Ontology (GO) terms was conducted using the online Gene Set Enrichment Analysis tool (https://www.gsea-msigdb.org/gsea/msigdb/annotate.jsp), which determines whether an *a priori* defined gene set shows statistically significant, concordant differences between two biological states. Using an FDR q-value < 0.05, we identified key biological pathways among the bi-directionally replicated genes associated with serum IgE concentration.

## Replication

### Study populations

Details of the replication studies (the Childhood Asthma Management Program (CAMP)^18–20^ and the Genetic Epidemiology of Asthma in Costa Rica Study (GACRS))^21^ have been described previously, including the assessment of IgE levels and gene expression profiling^20,22^. CAMP samples are from post-trial long-term follow-up blood draws. Written parental consent and child’s assent were obtained, and the study protocol was approved by the Institutional Review Boards at Hospital Nacional de Ninos (San Jose, Costa Rica) and Brigham and Women’s Hospital (Boston, MA).

### Statistical analysis

In both CAMP and GACRS, independent generalized linear regression models were run to test the association between each gene probe and log_10_transformed IgE concentration as aw continuous variable, using the “glmwrapper” function from the iCheck package with adjustment for age, sex, and the first two principal components. The Benjamini-Hochberg method was specified to control the false discovery rate with the q-value set to 0.05. The final dataset in GACRS included 25060 gene probes that passed QC from 326 subjects with available data and suitable samples; in CAMP 24972 gene probes from 610 participants were available. All probes measured in CAMP were also measured in GACRS.

### Meta-analysis

The results from CAMP and GACRS were meta-analyzed using the inverse normal method to combine p-values from the R package metaRNASeq^23^. Analyses were weighted according to the study size.

## Results

### FHS discovery TWAS of IgE levels

Clinical characteristics of FHS participants (mean age = 55 years; 54% women) and the replication cohorts (mean age = 20 and 9 years; 37% and 43% women in CAMP and GACRS, respectively) are presented in Supplementary Table 1. In FHS participants, among 17,873 mRNA gene-level transcripts that were available for analysis, 216 were associated with total IgE concentration at FDR< 0.05 (Supplementary Table 2) and 91 were significant at Bonferroni-corrected p-value threshold of p< 2.80×10^−6^ (0.05/17,873). The top thirty genes associated with serum IgE concentration are presented in Table 1. A volcano plot shows that the vast majority of genes at FDR< 0.05 (87.5% or 189/216) had expression levels that were positively associated with IgE (Figure 4).

**Table 1.** Top thirty genes associated with total IgE levels in the FHS.

| Gene Symbol | Chr | Beta | SE | P-Value | FDR Value |
| --- | --- | --- | --- | --- | --- |
| IL5RA | 3 | 0.144 | 0.011 | 1.88E-40 | 3.37E-36 |
| SLC29A1 | 6 | 0.085 | 0.007 | 3.23E-34 | 2.88E-30 |
| CLC | 19 | 0.172 | 0.014 | 3.60E-32 | 2.15E-28 |
| IL1RL1 | 2 | 0.131 | 0.011 | 6.41E-31 | 2.87E-27 |
| EMR1 | 19 | 0.111 | 0.010 | 1.13E-28 | 4.04E-25 |
| HRH4 | 18 | 0.118 | 0.011 | 8.13E-26 | 2.42E-22 |
| DACH1 | 13 | 0.050 | 0.005 | 4.26E-25 | 1.09E-21 |
| CCR4 | 3 | 0.084 | 0.008 | 1.04E-23 | 2.32E-20 |
| TEC | 4 | 0.067 | 0.007 | 1.31E-22 | 2.46E-19 |
| SYNE1 | 6 | 0.068 | 0.007 | 1.38E-22 | 2.46E-19 |
| ADORA3 | 1 | 0.061 | 0.006 | 1.32E-21 | 2.15E-18 |
| ALOX15 | 17 | 0.093 | 0.010 | 7.64E-21 | 1.14E-17 |
| CYSLTR2 | 13 | 0.089 | 0.010 | 4.43E-20 | 6.10E-17 |
| SMPD3 | 16 | 0.039 | 0.004 | 5.92E-20 | 7.55E-17 |
| IKZF2 | 2 | 0.058 | 0.007 | 3.18E-18 | 3.79E-15 |
| PRSS33 | 16 | 0.028 | 0.003 | 5.30E-18 | 5.92E-15 |
| PDE4D | 5 | 0.037 | 0.004 | 7.99E-18 | 8.40E-15 |
| CAT | 11 | 0.066 | 0.008 | 4.86E-17 | 4.82E-14 |
| SIGLEC8 | 19 | 0.042 | 0.005 | 1.52E-15 | 1.43E-12 |
| IDO1 | 8 | 0.070 | 0.009 | 3.20E-15 | 2.86E-12 |
| C2orf46 | 2 | 0.058 | 0.007 | 8.65E-15 | 7.37E-12 |
| VSTM1 | 19 | 0.087 | 0.011 | 5.22E-14 | 4.24E-11 |
| CD200R1 | 3 | 0.047 | 0.006 | 4.84E-13 | 3.76E-10 |
| ARHGAP10 | 4 | 0.033 | 0.005 | 7.98E-13 | 5.94E-10 |
| CCR3 | 3 | 0.070 | 0.010 | 2.38E-12 | 1.67E-09 |
| CEBPE | 14 | 0.048 | 0.007 | 2.43E-12 | 1.67E-09 |
| GPR114 | 16 | 0.023 | 0.003 | 5.64E-12 | 3.73E-09 |
| ANXA1 | 9 | 0.043 | 0.006 | 7.19E-12 | 4.59E-09 |
| C15orf43 | 15 | 0.037 | 0.005 | 8.92E-12 | 5.50E-09 |
| CAMK1 | 3 | 0.024 | 0.004 | 1.50E-11 | 8.95E-09 |

After adjusting for eosinophil count (Supplementary Table 3), fewer significant genes were identified (12 genes at FDR< 0.05, and six at Bonferroni-corrected p< 2.80×10^−6^). The attenuation of association is because eosinophil count was correlated with IgE level (R = 0.24, p< 1×10^−16^). There was concordance of effect estimates (betas) for the IgE-gene expression results with versus without adjustment for eosinophils (R = 0.46, p< 1×10^−16^; Supplementary Figure 1).

### Bi-directional Replication

Out of 216 unique transcripts at FDR< 0.05 from discovery in FHS, 59 unique transcripts replicated in the meta-analyzed results from GACRS and CAMP (Supplementary Table 4). We defined replication as genes at p-value < 2.44×10^−4^ (0.05/205), as only 205 of the 216 significant genes in FHS were available for analysis in the replication cohorts. Forest plots of the top ten genes in this replicated gene set are provided in Supplementary Figure 2.

We performed reverse replication with the meta-analysis of GACRS/CAMP as the discovery set and FHS as the replication set. From the meta-analysis of GACRS/CAMP, we identified 135 unique transcripts associated with total IgE levels at FDR< 0.05. Among these, 114 transcripts mapping to 112 unique genes were available in FHS (*TRERF1* and *ACOT11* were each linked to two separate transcripts). We defined replication as p< 4.39×10^−4^ (0.05/114); all 114 significant transcripts from discovery in GACRS/CAMP replicated in FHS (Supplementary Table 5). Furthermore, all 59 genes that replicated in GACRS/CAMP based on FHS discovery were within the 114 replicated gene set using GACRS/CAMP as discovery—i.e. 59 genes demonstrated bi-directional replication, demonstrating the robustness of association signals.

### Gene Ontology

Using the 216 FDR-signficant genes from FHS discovery, we conducted gene ontology to identify key biological pathways among the top genes differentially expressed in relation to total IgE levels (Table 2). Multiple genes from this gene set were associated with pathways involved in inflammation and other immune system responses.

**Table 2.** Gene ontology defned biological processes associated with total IgE levels in the FDR-significant transcripts in the FHS cohort (n = 216).

| GO biological process | # Genes in GO group | P-Value | FDR Value |
| --- | --- | --- | --- |
| Regulation of Immune System Process | 40 | 6.69E-15 | 1.98E-11 |
| Defense Response | 41 | 8.10E-15 | 1.98E-11 |
| Inflammatory Response | 27 | 3.58E-14 | 5.82E-11 |
| Leukocyte Migration | 21 | 1.21E-12 | 1.47E-09 |
| Neurogenesis | 35 | 8.15E-12 | 7.96E-09 |
| Immune System Development | 27 | 1.42E-11 | 1.26E-08 |
| Leukocyte Proliferation | 14 | 2.0E-09 | 6.98E-07 |
| Cytokine Mediated Signaling Pathway | 21 | 4.84E-09 | 1.57E-06 |
| Response to Cytokine | 26 | 5.35E-09 | 1.67E-06 |
| Reactome Signaling by Interleukins | 16 | 9.39E-09 | 2.58E-06 |
| Cytokine Production | 20 | 3.62E-08 | 7.62E-06 |
| Lymphocyte Activation | 19 | 4.2E-08 | 8.54E-06 |
| Neuron Differentiation | 26 | 5.97E-08 | 1.17E-05 |
| Positive Regulation of Hemopoiesis | 10 | 1.23E-07 | 2.03E-05 |
| Interleukin 5 Production | 5 | 1.28E-07 | 2.08E-05 |
| Myeloid Cell Homeostasis | 9 | 1.61E-07 | 2.54E-05 |
| Leukocyte Cell Cell Adhesion | 12 | 6.28E-07 | 7.96E-05 |
| Immune Effector Process | 23 | 8.35E-07 | 9.93E-05 |
| Myeloid Cell Differentiation | 13 | 8.44E-07 | 9.93E-05 |
| Neuron Development | 21 | 1.43E-06 | 1.57E-04 |
| Granulocyte Migration | 8 | 1.49E-06 | 1.62E-04 |
| Leukocyte Differentiation | 14 | 1.65E-06 | 1.77E-04 |

Secondary gene ontology analysis was performed on the 59 genes that bi-directionally replicated between FHS and GACRS/CAMP. Similar to the larger FHS discovery gene set, multiple genes from the bidirectionally replicated gene set were implicated in pathways related to the immune response (Supplementary Table 6).

### Mendelian Randomization for total IgE levels

A Manhattan plot and a Q-Q plot (lambda 1.017) displaying the updated FHS IgE GWAS results are provided in Figures 2 and 3, respectively. A list of significant SNPs (p< 5×10^−8^) from the updated IgE GWAS is reported in Supplementary Table 7. Among the 216 FDR-significant genes identified in FHS, 185 genes had suitable *cis-*eQTLs for the MR analysis. We conducted bi-directional MR to test causal relations between expression levels of the 185 genes and circulating IgE levels. We identified four genes—*CLC, CCDC21, S100A13*, and *GCNT1*—as putatively causal for IgE at P_mRNA→IgE_ < 0.05 using the top *cis-*eQTL for each gene as an instrument variable (Table 3).

**Figure 2.**
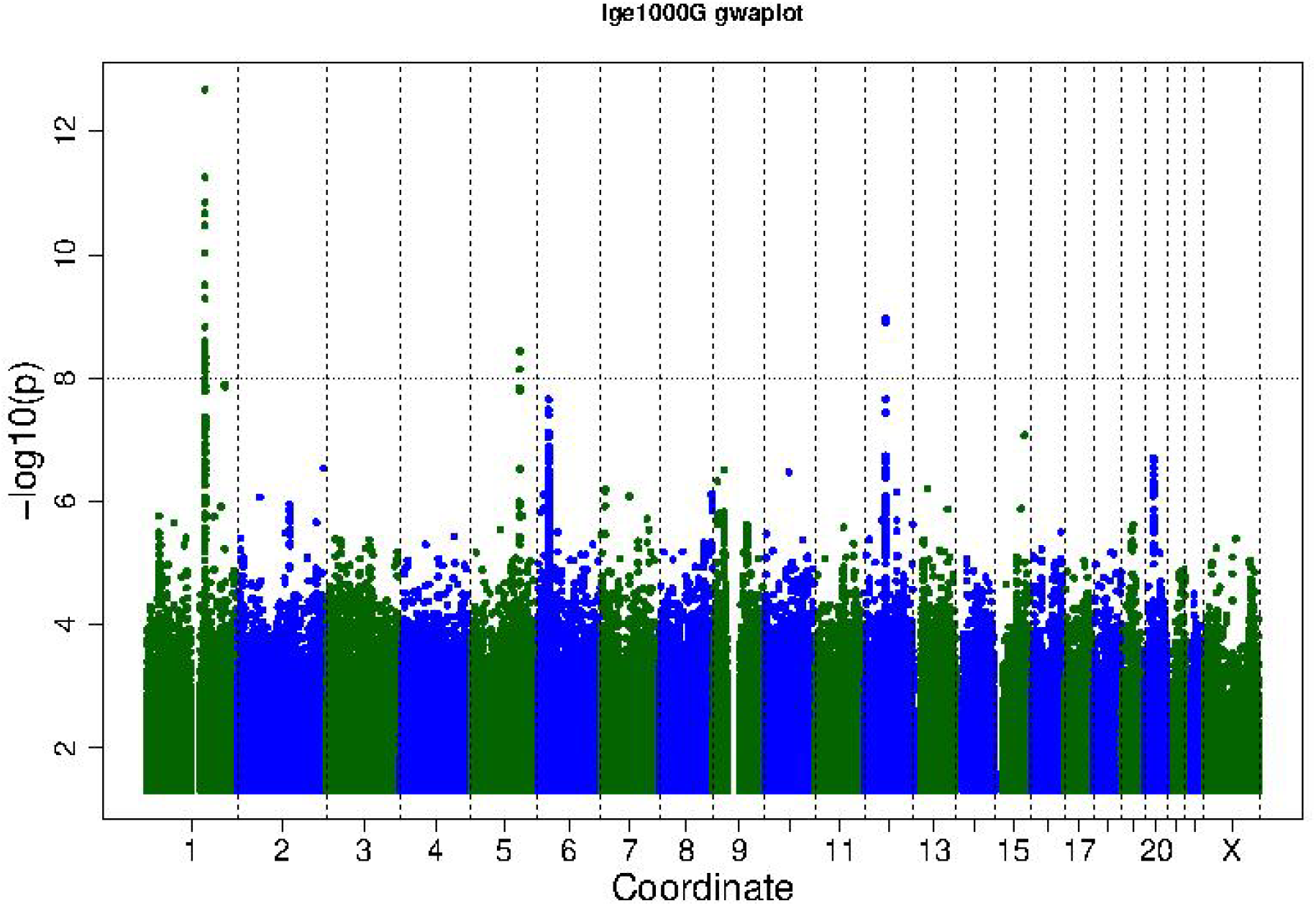
Manhattan plot of FHS GWAS results. The horizontal line shows the threshold for genome-wide significance (P-value < 5×10^−8^).

**Figure 3.**
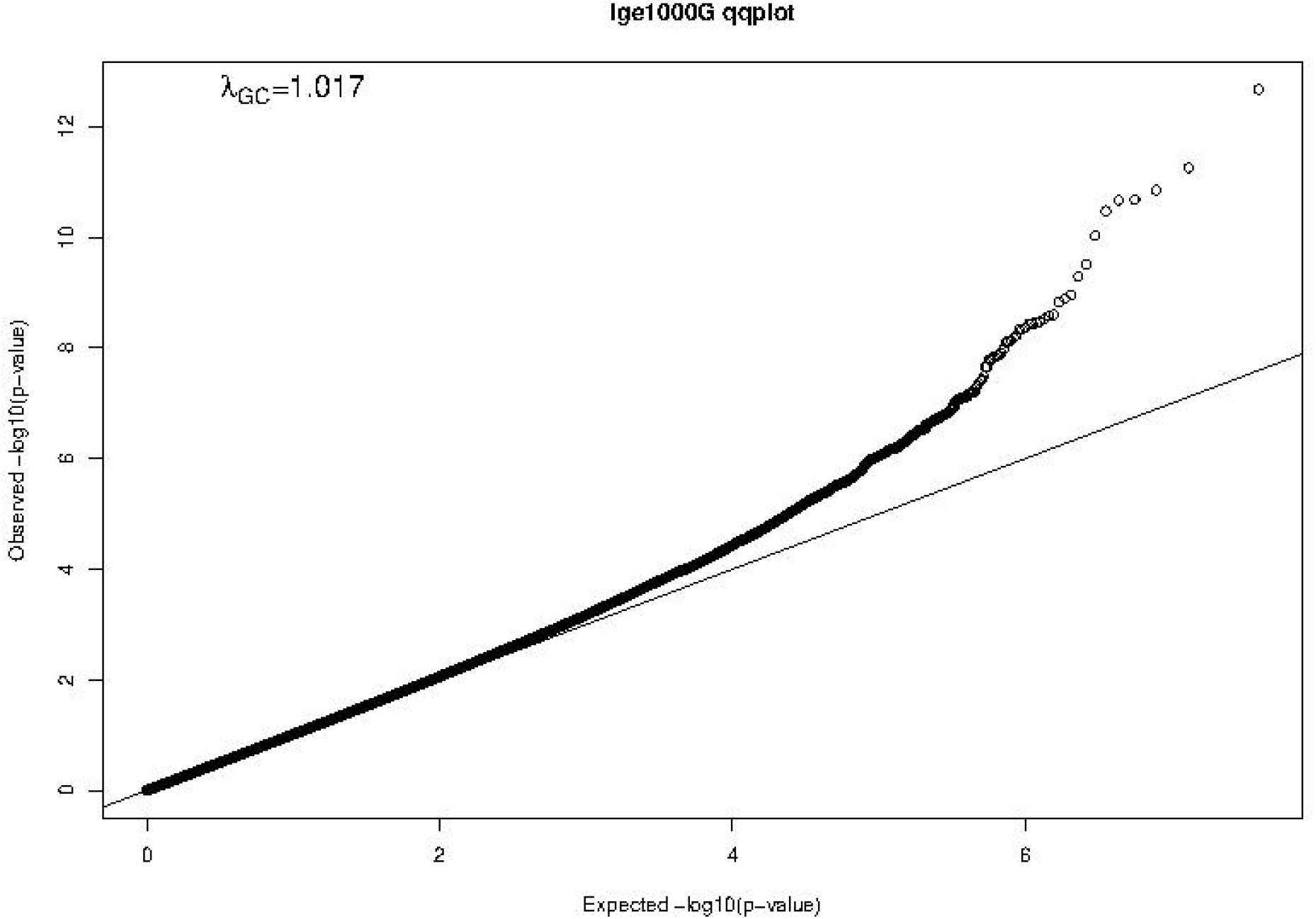
Q-Q plot of FHS GWAS results. The Q-Q plot shows observed vs. expected -log10(P-values)
of the GWAS results. The straight line represents the SNP distribution under the null hypothesis.

**Figure 4.**
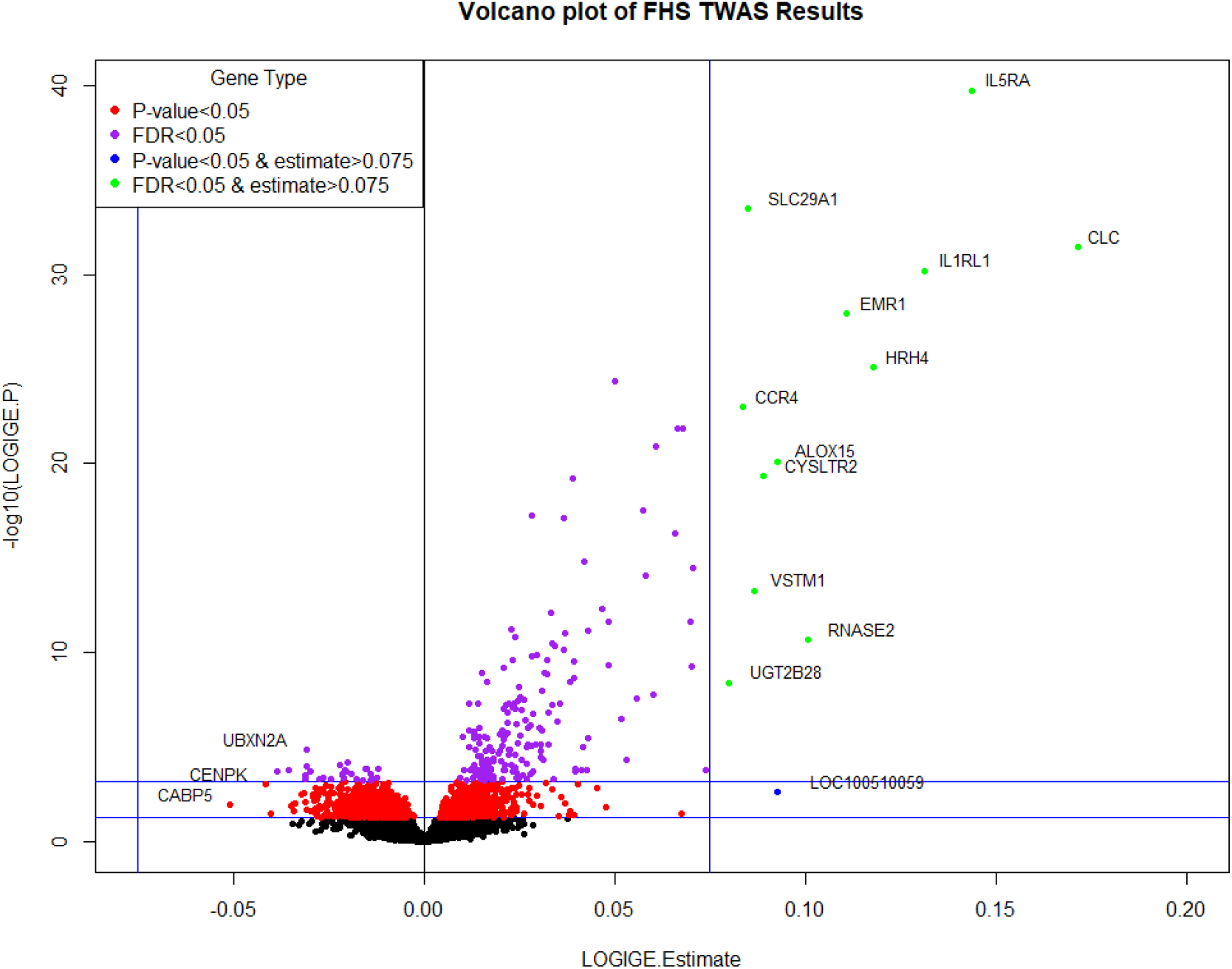
Volcano plot of FHS TWAS results. P-value < 0.05 significance threshold is the lower line and P-value < 6.09×10^−4^ (which corresponds to an FDR < 0.05) is the upper line.

**Table 3.** Bi-directional MR results for genes putatively causal for IgE levels at p< 0.05 (n = 4).

| Gene Symbol | SNP | Chr | Beta | SE | P-Value | Beta | SE | P-Value |
| --- | --- | --- | --- | --- | --- | --- | --- | --- |
| | | | (mRNA→IgE) | (mRNA→IgE) | (mRNA→IgE) | (IgE→mRNA) | (IgE→mRNA) | (IgE→mRNA) |
| CCDC21 | rs869683 | 1 | 0.571 | 0.197 | 3.67E-03 | -0.005 | 0.024 | 8.46E-01 |
| S100A13 | rs9661993 | 1 | -0.166 | 0.062 | 7.35E-03 | -0.097 | 0.057 | 9.15E-02 |
| GCNT1 | rs11144929 | 9 | 1.503 | 0.606 | 1.32E-02 | -0.018 | 0.03 | 5.62E-01 |
| CLC | rs17709471 | 19 | 0.467 | 0.198 | 1.80E-02 | -0.061 | 0.082 | 4.58E-01 |

Additionally, we performed reverse MR using the top six SNPs from IgE GWAS combined as a polygenic risk score to test if IgE level affected gene expression levels. None of the four genes from forward MR were significant in reverse MR (P_IgE→mRNA_ ≥ 0.05) (Table 3), suggesting a stronger likelihood that gene expression drives changes in IgE levels rather than IgE levels driving gene expression.

### Mendelian Randomization for IgE-related diseases: asthma and allergic diseases

We conducted two-sample MR tests to infer a causal relation between IgE-related gene expression and IgE-related diseases, specifically asthma and allergic diseases. We identified 70 genes that were putatively causal for asthma and 71 genes that were putatively causal for allergic diseases at a Bonferroni-corrected p-value threshold of p< 2.70×10^−4^ (0.05/185) (Table 4, Supplementary Table 8). In comparing the MR results of asthma to those of allergic diseases, the vast majority of putatively causal genes (68) overlapped, which is to be expected given that asthma and allergic diseases are IgE-related (Table 4).

**Table 4.** MR results for genes putatively causal for asthma at Bonferroni-corrected p< 2.70×10^−4^ (n = 70).

| Gene Symbol | Transcript # | Method | nsnp | Beta | SE | P-Value |
| --- | --- | --- | --- | --- | --- | --- |
| GPI | 3829687 | Wald ratio | 1 | 23.67 | 0.62 | 0.00E+00 |
| PGD | 2319802 | Wald ratio | 1 | 70.20 | 1.08 | 0.00E+00 |
| NDFIP2 | 3495076 | Wald ratio | 1 | 76.43 | 1.01 | 0.00E+00 |
| CYSLTR2 | 3489138 | Wald ratio | 1 | 21.71 | 0.30 | 0.00E+00 |
| GPR114 | 3662774 | Wald ratio | 1 | -60.23 | 0.94 | 0.00E+00 |
| CPT1A | 3379644 | Wald ratio | 1 | 34.88 | 0.66 | 0.00E+00 |
| ZNF610 | 3840164 | Wald ratio | 1 | -51.06 | 0.72 | 0.00E+00 |
| ABTB2 | 3368940 | Wald ratio | 1 | 48.78 | 0.67 | 0.00E+00 |
| CD9 | 3402315 | Wald ratio | 1 | 26.12 | 0.55 | 0.00E+00 |
| STK17A | 2999485 | Wald ratio | 1 | 42.04 | 0.71 | 0.00E+00 |
| SMPD3 | 3696317 | Wald ratio | 1 | -59.35 | 1.01 | 0.00E+00 |
| GCNT1 | 3175494 | Wald ratio | 1 | 58.12 | 0.84 | 0.00E+00 |
| CCR4 | 2616131 | Wald ratio | 1 | 20.37 | 0.32 | 0.00E+00 |
| PRF1 | 3293435 | Wald ratio | 1 | 40.38 | 0.93 | 0.00E+00 |
| ATP8B2 | 2360206 | Wald ratio | 1 | 47.85 | 0.71 | 0.00E+00 |
| ZMYND11 | 3231389 | Wald ratio | 1 | -71.58 | 1.00 | 0.00E+00 |
| OLIG1 | 3918429 | Wald ratio | 1 | 96.66 | 1.41 | 0.00E+00 |
| FBP1 | 3215570 | Wald ratio | 1 | -62.42 | 1.16 | 0.00E+00 |
| ORM2 | 3186137 | Wald ratio | 1 | -29.95 | 0.38 | 0.00E+00 |
| SAMSN1 | 3925473 | Wald ratio | 1 | 51.39 | 0.82 | 0.00E+00 |
| FCRL2 | 2439052 | Wald ratio | 1 | -52.09 | 0.83 | 0.00E+00 |
| CCDC86 | 3332548 | Wald ratio | 1 | 58.53 | 0.80 | 0.00E+00 |
| KIT | 2727587 | Wald ratio | 1 | 86.75 | 1.15 | 0.00E+00 |
| PLD3 | 3833443 | Wald ratio | 1 | 40.25 | 0.87 | 0.00E+00 |
| BHLHE40 | 2608725 | Wald ratio | 1 | -24.02 | 0.47 | 0.00E+00 |
| THUMPD1 | 3683783 | Wald ratio | 1 | 31.31 | 0.42 | 0.00E+00 |
| CLINT1 | 2883609 | Wald ratio | 1 | 59.72 | 0.82 | 0.00E+00 |
| IL5RA | 2660617 | Wald ratio | 1 | 44.95 | 0.62 | 0.00E+00 |
| IL2RA | 3275729 | Wald ratio | 1 | 28.73 | 0.40 | 0.00E+00 |
| SRGAP3 | 2662087 | Wald ratio | 1 | 79.94 | 1.07 | 0.00E+00 |
| NUPL1 | 3482219 | Wald ratio | 1 | 53.59 | 1.27 | 0.00E+00 |
| NUP93 | 3662265 | Wald ratio | 1 | 53.06 | 0.66 | 0.00E+00 |
| IL17RB | 2624565 | Wald ratio | 1 | -33.81 | 0.45 | 0.00E+00 |
| CRIP1 | 3554851 | Wald ratio | 1 | 28.22 | 0.43 | 0.00E+00 |
| GPR137B | 2386747 | Wald ratio | 1 | -65.16 | 0.85 | 0.00E+00 |
| ID2 | 2468622 | Wald ratio | 1 | -38.70 | 0.49 | 0.00E+00 |
| CLCNKB | 2322264 | Wald ratio | 1 | 68.19 | 0.93 | 0.00E+00 |
| PLAC4 | 3932917 | Wald ratio | 1 | -15.40 | 0.21 | 0.00E+00 |
| SYNE1 | 2979871 | Wald ratio | 1 | -45.92 | 1.08 | 0.00E+00 |
| FCRLA | 2363852 | Wald ratio | 1 | -66.59 | 0.70 | 0.00E+00 |
| COBLL1 | 2584787 | Wald ratio | 1 | 84.94 | 1.28 | 0.00E+00 |
| INPP5A | 3272205 | Wald ratio | 1 | -22.45 | 0.50 | 0.00E+00 |
| SEMA7A | 3632907 | Wald ratio | 1 | -49.69 | 1.14 | 0.00E+00 |
| ADORA3 | 2427981 | Inverse variance weighted | 2 | 19.38 | 0.43 | 0.00E+00 |
| GATA3 | 3234277 | Wald ratio | 1 | 42.41 | 0.58 | 0.00E+00 |
| PMP22 | 3746574 | Inverse variance weighted | 2 | 30.48 | 0.82 | 5.27E-303 |
| P4HA1 | 3294159 | Wald ratio | 1 | 10.88 | 0.30 | 1.23E-288 |
| KLHL6 | 2708066 | Wald ratio | 1 | -21.75 | 0.65 | 5.52E-244 |
| PPM1L | 2650393 | Wald ratio | 1 | -11.22 | 0.35 | 5.00E-224 |
| GRB10 | 3050462 | Wald ratio | 1 | 28.66 | 0.94 | 5.03E-204 |
| CDK15 | 2522916 | Wald ratio | 1 | 10.88 | 0.39 | 5.54E-175 |
| BRI3BP | 3436544 | Wald ratio | 1 | 31.48 | 1.14 | 5.51E-169 |
| SEMA5A | 2847967 | Wald ratio | 1 | 36.84 | 1.44 | 3.84E-145 |
| HRASLS2 | 3376512 | Wald ratio | 1 | 10.69 | 0.43 | 7.30E-139 |
| PDE8A | 3606034 | Wald ratio | 1 | 23.80 | 1.16 | 2.96E-93 |
| CD63 | 3457160 | Inverse variance weighted | 2 | -80.22 | 5.03 | 3.30E-57 |
| VKORC1L1 | 3005280 | Wald ratio | 1 | 22.91 | 1.44 | 8.49E-57 |
| SLC35D1 | 2417095 | Inverse variance weighted | 3 | 14.53 | 1.87 | 8.13E-15 |
| FMNL3 | 3454006 | Inverse variance weighted | 2 | -37.28 | 5.70 | 6.10E-11 |
| TNIK | 2705266 | Inverse variance weighted | 3 | -64.09 | 9.84 | 7.23E-11 |
| VSTM1 | 3870449 | Inverse variance weighted | 3 | -6.97 | 1.17 | 2.49E-09 |
| CCL23 | 3753985 | Inverse variance weighted | 2 | -36.91 | 7.31 | 4.35E-07 |
| VLDLR | 3160175 | Inverse variance weighted | 5 | 33.87 | 6.73 | 4.76E-07 |
| TNFRSF9 | 2395146 | Inverse variance weighted | 2 | -12.54 | 2.60 | 1.41E-06 |
| ABCC1 | 3649890 | Inverse variance weighted | 2 | -44.86 | 10.32 | 1.39E-05 |
| KLF6 | 3274361 | Inverse variance weighted | 2 | -54.40 | 13.38 | 4.79E-05 |
| CASP3 | 2796484 | Inverse variance weighted | 2 | -11.03 | 2.74 | 5.69E-05 |
| TEC | 2768396 | Inverse variance weighted | 3 | -22.47 | 5.73 | 8.74E-05 |
| GZMB | 3558375 | Inverse variance weighted | 2 | -13.92 | 3.61 | 1.17E-04 |
| INPP1 | 2520113 | Inverse variance weighted | 2 | -25.81 | 6.83 | 1.58E-04 |

*GCNT1*, a putatively causal gene for IgE concentration as implicated in our MR analysis of gene expression in relation to IgE levels (beta = 1.503, p = 1.32×10^−2^; Table 3), is also one of the top results in the MR analysis of expression in relation to asthma and allergic diseases (beta = 58.12, p< 1×10^−400^ and beta = 58.88, p< 1×10^−400^, respectively; Table 4, Supplementary Table 8).

## Discussion

A thorough understanding of the molecular mechanisms underlying the regulation of IgE is essential for developing new therapies for asthma and other IgE-mediated diseases, such as allergic rhinitis, atopic dermatitis, and food allergies. To the best of our knowledge, this is the first large-scale TWAS study of total IgE levels that uses MR to infer causal relations between gene expression and IgE levels. In this study, we identified a transcriptomic signature of IgE consisting of 216 FDR-significant genes from discovery in FHS. Gene ontology analysis of this gene set shows that many of these IgE-related genes are enriched in key pathways related to regulation of immune system processes, defense response, and inflammatory response.

MR analysis revealed four genes (*CLC, CCDC21, S100A13*, and *GCNT1*) as nominally significant (P_mRNA→IgE_ < 0.05) causal regulators of IgE concentration, suggesting that individual gene transcripts that are associated with IgE concentration likely contribute causally to IgE regulation. Among these is *CLC* (Charcot-Leydon crystal galectin), which is overexpressed in eosinophils that are stimulated following binding of IgE^24^. Prior studies have identified increased CLC protein levels in induced sputum as a surrogate biomarker of eosinophilic airway inflammation in asthma^25^. Another recent study used a humanized mouse model of asthma to demonstrate that administration of CLC protein with house dust mites (HDM) increased human IgE synthesis compared to when HDMs were administered alone. After developing antibodies that specifically bind and dissolve CLC crystals, it was found that antibiotic treatment completely neutralized the inflammatory effect of CLCs and the ability of CLCs to enhance IgE synthesis^12,26^. The strong association of the protein encoded by *CLC* with IgE concentration, revealed by our TWAS and MR analysis, highlights *CLC* as a key gene and attractive therapeutic target. While this association does not persist after adjusting for eosinophil count, it is likely because the mechanisms by which *CLC* SNPs and *CLC* expression influence IgE concentration—and presumably asthma and allergic diseases—are mediated by eosinophils; thus, adjusting for eosinophils would be expected to mask these associations and may be an overadjustment.

An emerging area of interest in immunology in recent years is the effects on immunity and disease susceptibility of glycosylation of lipid or protein molecules by glycans such as *GCNT1* (glucosaminyl (N-acetyl) transferase 1)^27^. *GCNT1* is a glycosyltransferase involved in pathways related to metabolism of proteins, and it has several functions involved in immune response. One recent study demonstrated that the protein product of *GCNT1*, core 2 ß1,6-*N*-acetylglucosaminyltransferase-I (C2GlcNAcT-I), is necessary not only for the synthesis of P-selectin ligands in neutrophils and T helper 1 (Th1) cells but also for the homing of Th1 cells into sites of inflammation^28^. Additional roles of *GCNT1* include partially controlling lymphocyte trafficking into lymph nodes and regulating B cell differentiation via formation and extension of core 2 O-glycans^29,30^. These functions are critical to understanding the relations of *GCNT1* to IgE concentration given that B cells produce IgE.

Interestingly, a recent knockout study found that *GCNT1* deficient mice have neutrophilia and increased susceptibility to tuberculosis infection. The increased susceptibility of *GCNT1* deficient mice to infection was largely driven by exacerbated neutrophil counts, which led to lung lesions, inflammation, and other pathologic features in the lungs of affected mice^31^. This link between *GCNT1* and neutrophilia is relevant to studying the regulation of IgE as other studies have shown elevated serum IgE levels to be associated with neutrophilic asthma^32^. Therefore, it is possible that a deficiency, or more broadly an alteration, in *GCNT1* levels may be linked with elevated IgE levels; additional functional studies are warranted to explore the relationship between *GCNT1* and serum IgE concentration. Given that there is no previously published causal association between *GCNT1* and IgE concentration and that *GCNT1* appears to play a role in immune processes such as inflammatory Th1 homing, lymphocyte trafficking, and B cell differentiation, *GCNT1* represents a highly promising therapeutic target for the treatment and prevention of asthma and IgE-related diseases.

Two other nominally significant genes implicated in MR testing—*CCDC21* and *S100A13*—have no known mechanistic association with serum IgE concentration. *CCDC21* encodes a protein (centrosomal protein 85) that belongs to the centrosome-associated family of proteins. *S100A13* is a calcium binding gene that encodes for a protein (S100 calcium binding protein A13) belonging to the S100 family of proteins that are involved in a broad range of intracellular and extracellular functions. Extracellular S100 proteins often play crucial roles in regulating immune homeostasis and inflammation^33^. By interacting with cell surface receptors such as RAGE (receptor for advanced glycation end products) in response to cell stress or inflammation, S100 proteins can activate intracellular signaling pathways that induce production of pro-inflammatory cytokines and lead to the migration of neutrophils, monocytes, and macrophages^33^. Various extracellular S100 proteins have been associated with the pathogenesis of inflammatory diseases such as allergy. For example, multiple anti-allergic drugs such as amlexanox, cromolyn, and tranilast have been shown to bind S100A13 and block downstream RAGE signaling^33^. While *CCDC21* and *S100A13* have not previously been shown to have roles in IgE regulation, our MR tests implicate them as potentially novel biomarkers or therapeutic targets.

In MR analyses of IgE-related diseases, we identified an IgE-associated gene expression signature that is “putatively” causal for asthma. Similar MR results for allergic diseases serve as further confirmation of our MR results of asthma. The identification of *GCNT1* as a causal gene for IgE concentration, asthma, and allergic diseases provides additional support for our hypothesis that IgE-associated gene expression changes impact IgE regulation and play a role in multiple IgE-related diseases. Based on our finding of a putatively causal role of *GCNT1* in IgE regulation and in asthma and allergic diseases, we hypothesize that *GCNT1* and the other IgE-associated genes identified in this study are related to the pathobiology of IgE-related diseases, including asthma and allergic diseases, and that they represent compelling therapeutic targets for treatment and prevention of these disorders.

There are several limitations to our study. First, gene expression was measured in whole blood, which may not be representative of tissue-specific effects. Second, the serum IgE concentration and gene expression measurements were obtained approximately six years apart in FHS participants. This time difference may bias our results toward the null. Third, there are significant differences in the age and IgE concentration of the FHS participants compared to those of GACRS/CAMP. The average age of the FHS study participants was 55 years, which was significantly higher than the average age of GACRS (9 years) and CAMP (20 years) participants. There was no significant difference in serum IgE concentration between the GACRS and CAMP cohorts, despite the ten-year age difference; however, the IgE levels of the GACRS and CAMP cohorts were considerably higher than those of FHS (log_10_IgE levels 2.5 and 2.5 vs. 1.52). This is likely due to the fact that all the participants in GACRS and CAMP had asthma, which is associated with elevated IgE concentration.

Of the 216 IgE-associated transcripts (FDR< 0.05) in FHS discovery, 59 genes bi-directionally replicated between the FHS and the GACRS/CAMP. This high-degree of replication is notable given the previously described differences in cohort study populations.

We performed a TWAS of IgE and then probed the directional relations between IgE and gene expression, which identified four genes as causally associated with IgE levels. *CLC* is a well-documented gene with known associations with eosinophils and IgE; *CCDC21* and *S100A13* do not yet have well-understood associations with IgE and represent novel findings. Given its myriad of roles in the regulation of the immune response, *GCNT1* is a particularly attractive potential drug target given that in addition to its putatively causal relation to IgE levels it also was causal for asthma and allergic diseases. Our findings build upon prior knowledge of IgE regulation and provide a deeper understanding of the underlying molecular mechanisms. The IgE-associated genes that we identified—particularly those implicated in MR testing—can be explored as promising therapeutic targets for asthma and IgE-related diseases.

## Data Availability

The datasets generated during and/or analysed during the current study are available from the corresponding author on reasonable request.

## Acknowledgments

The Framingham Heart Study is funded by National Institutes of Health contract N01-HC-25195 and HHSN268201500001I. The work in this project was funded by the Division of Intramural Research, National Heart, Lung, and Blood Institute, National Institutes of Health, Bethesda, MD (D. Levy, Principal Investigator).

## Disclaimer

The views expressed in this manuscript are those of the authors and do not necessarily represent the views of the National Heart, Lung, and Blood Institute; the National Institutes of Health; or the U.S. Department of Health and Human Services.

## References

1. Oettgen, H.C. Fifty years later: Emerging functions of IgE antibodies in host defense, immune regulation, and allergic diseases. The Journal of allergy and clinical immunology 137, 1631–1645 (2016).

2. Kelly, B.T. & Grayson, M.H. Immunoglobulin E, what is it good for? Annals of allergy, asthma & immunology: official publication of the American College of Allergy, Asthma, & Immunology 116, 183–187 (2016).

3. Mukai, K., Tsai, M., Starkl, P., Marichal, T. & Galli, S.J. IgE and mast cells in host defense against parasites and venoms. Seminars in Immunopathology 38, 581–603 (2016).

4. Hu, J. et al. Anti-IgE therapy for IgE-mediated allergic diseases: from neutralizing IgE antibodies to eliminating IgE(+) B cells. Clinical and translational allergy 8, 27–27 (2018).

5. Levin, A.M. et al. A meta-analysis of genome-wide association studies for serum total IgE in diverse study populations. The Journal of allergy and clinical immunology 131, 1176–1184 (2013).

6. Yatagai, Y. et al. Genome-wide association study for levels of total serum IgE identifies HLA-C in a Japanese population. PloS one 8, e80941-e80941 (2013).

7. Moffatt, M.F. et al. A large-scale, consortium-based genomewide association study of asthma. The New England journal of medicine 363, 1211–1221 (2010).

8. Granada, M. et al. A genome-wide association study of plasma total IgE concentrations in the Framingham Heart Study. The Journal of allergy and clinical immunology 129, 840–845.e21 (2012).

9. Liang, L. et al. An epigenome-wide association study of total serum immunoglobulin E concentration. Nature 520, 670–674 (2015).

10. Weidinger, S. et al. Genome-wide scan on total serum IgE levels identifies FCER1A as novel susceptibility locus. PLoS genetics 4, e1000166-e1000166 (2008).

11. Everson, T.M. et al. DNA methylation loci associated with atopy and high serum IgE: a genome-wide application of recursive Random Forest feature selection. Genome medicine 7, 89–89 (2015).

12. Virkud, Y.V. et al. Novel eosinophilic gene expression networks associated with IgE in two distinct asthma populations. Clin Exp Allergy 48, 1654–1664 (2018).

13. Zhu, Z. et al. A genome-wide cross-trait analysis from UK Biobank highlights the shared genetic architecture of asthma and allergic diseases. Nat Genet 50, 857–864 (2018).

14. Dawber, T.R., Meadors, G.F. & Moore, F.E., Jr. Epidemiological approaches to heart disease: the Framingham Study. Am J Public Health Nations Health 41, 279–81 (1951).

15. Yunginger, J.W. et al. Quantitative IgE antibody assays in allergic diseases. J Allergy Clin Immunol 105, 1077–84 (2000).

16. Joehanes, R. et al. Integrated genome-wide analysis of expression quantitative trait loci aids interpretation of genomic association studies. Genome biology 18, 16 (2017).

17. Joehanes, R. et al. Integrated genome-wide analysis of expression quantitative trait loci aids interpretation of genomic association studies. Genome Biol 18, 16 (2017).

18. The Childhood Asthma Management Program (CAMP): design, rationale, and methods. Childhood Asthma Management Program Research Group. Control Clin Trials 20, 91–120 (1999).

19. Recruitment of participants in the childhood Asthma Management Program (CAMP). I. Description of methods: Childhood Asthma Management Program Research Group. J Asthma 36, 217–37 (1999).

20. Croteau-Chonka, D.C. et al. Gene Expression Profiling in Blood Provides Reproducible Molecular Insights into Asthma Control. Am J Respir Crit Care Med 195, 179–188 (2017).

21. Kelly, R.S. et al. Metabolomic profiling of lung function in Costa-Rican children with asthma. Biochim Biophys Acta Mol Basis Dis 1863, 1590–1595 (2017).

22. Bourgon, R., Gentleman, R. & Huber, W. Independent filtering increases detection power for high-throughput experiments. Proceedings of the National Academy of Sciences of the United States of America 107, 9546–9551 (2010).

23. Rau, A., Marot, G. & Jaffrezic, F. Differential meta-analysis of RNA-seq data from multiple studies. BMC Bioinformatics 15, 91 (2014).

24. Su, J. A. Brief History of Charcot-Leyden Crystal Protein/Galectin-10 Research. Molecules (Basel, Switzerland) 23, 2931 (2018).

25. Nyenhuis, S.M. et al. Charcot-Leyden crystal protein/galectin-10 is a surrogate biomarker of eosinophilic airway inflammation in asthma. Biomark Med 13, 715–724 (2019).

26. Persson, E.K. et al. Protein crystallization promotes type 2 immunity and is reversible by antibody treatment. Science 364(2019).

27. Lyons, J.J., Milner, J.D. & Rosenzweig, S.D. Glycans Instructing Immunity: The Emerging Role of Altered Glycosylation in Clinical Immunology. Frontiers in pediatrics 3, 54–54 (2015).

28. Mardahl, M. et al. Core 2 ß1,6-N-acetylglucosaminyltransferase-I, crucial for P-selectin ligand expression is controlled by a distal enhancer regulated by STAT4 and T-bet in CD4+ T helper cells 1. Mol Immunol 77, 132–40 (2016).

29. Veerman, K., Tardiveau, C., Martins, F., Coudert, J. & Girard, J.P. Single-Cell Analysis Reveals Heterogeneity of High Endothelial Venules and Different Regulation of Genes Controlling Lymphocyte Entry to Lymph Nodes. Cell Rep 26, 3116–3131.e5 (2019).

30. Giovannone, N. et al. Human B Cell Differentiation Is Characterized by Progressive Remodeling of O-Linked Glycans. Frontiers in immunology 9, 2857–2857 (2018).

31. Fonseca, K.L. et al. Deficiency in the glycosyltransferase Gcnt1 increases susceptibility to tuberculosis through a mechanism involving neutrophils. Mucosal Immunology (2020).

32. Bullone, M. et al. Elevated serum IgE, oral corticosteroid dependence and IL-17/22 expression in highly neutrophilic asthma. European Respiratory Journal 54, 1900068 (2019).

33. Xia, C., Braunstein, Z., Toomey, A.C., Zhong, J. & Rao, X. S100 Proteins As an Important Regulator of Macrophage Inflammation. Front Immunol 8, 1908 (2017).

